# Advanced neurological recovery translates into greater long-term functional independence after acute spinal cord injury

**DOI:** 10.1101/2020.09.01.20185413

**Authors:** Navid Khosravi-Hashemi, Rainer Abel, Lukas Grassner, Yorck-Bernhard Kalke, Doris Maier, Rüdiger Rupp, Norbert Weidner, Armin Curt, John K. Kramer

## Abstract

The absence of effective pharmacological interventions in acute traumatic spinal cord injury is a major problem in its management. A critical barrier in identifying such interventions lies in the vast heterogeneity of recovery profiles, which masks the potential efficacy of treatments in clinical trials. To determine the impact of temporal recovery profiles on long-term functional independence, we used EMSCI (European Multicenter Study about Spinal Cord Injury) data. Total motor scores from the International Standards for the Neurological Classification of Spinal Cord Injury (ISNCSCI) and the Spinal Cord Independence Measure (SCIM) were used to assess neurological and functional outcomes, respectively. We developed a classification method consisting of thresholding and unsupervised machine learning clustering and applied it to the total motor score profiles. Comparing SCIM scores between classes revealed that functional independence is significantly higher among patients displaying advanced neurological recovery profile. Our study suggests that the evaluation of temporal recovery profiles can provide novel insights in spinal cord injury clinical trials.

## INTRODUCTION

Acute traumatic spinal cord injury represents a significant life-event, characterized by motor, sensory, and autonomic deficits^1,2^. A major problem facing its management is the absence of pharmacological interventions that protect and/or repair the damaged cord. These are urgently needed to increase the amount of neurological and functional recovery, beyond that expected spontaneously^3^. A comprehensive understanding of spontaneous recovery and its heterogeneity will facilitate the discovery of such interventions.

Spontaneous neurological recovery is typically characterized by rapid gains in function in the initial days to weeks post-injury, followed by substantially slower progression before plateauing between 6 and 12 months^4^. This classical recovery profile has been described in numerous studies^5,6,7,8^, and is preserved on a reduced time-scale in animal models^9,10^. On the individual subject level, there is, however, a considerable degree of variability, with some persons recovering to a greater degree and/or much faster compared to others^11^.

While increasing the total amount of neurological function recovered represents an important goal of acute therapeutic interventions^12,13^, the value of advanced recovery is mostly unknown. In preclinical animal models, particularly in peripheral injuries but also after damage in the spinal cord, “advanced recovery” is a common effect of an intervention (e.g., electrical stimulation)^14,15,16^.

The current study aimed to determine the extent that advanced neurological recovery mattered during the transition from acute to chronic spinal cord injury. We hypothesized that individuals recovering neurological function at an early high pace would be more independent at 1-year post-injury compared to individuals who recovered slower at first but to a similar overall extent. To address this hypothesis, we applied an unbiased, machine learning approach to analyze motor recovery and functional independence in individuals with acute spinal cord injury.

## RESULTS

To investigate the relation between the profile of neurological recovery and long-term functional independence, we developed a method to classify patients based on their neurological recovery profile. We used the area under the curves (AUC) to distinguish “advanced” and “delayed” recovery profiles. In brief, we fit linear interpolation curves to motor scores over the first year post-injury, normalized the curves, calculated the AUC’s of the normalized curves (AUNC), and applied an unsupervised machine learning clustering approach called “K-means” ^17^. To test if this method was accurately estimating advanced and delayed recovery, we performed a simulation study. In this simulation, motor scores were generated randomly and ordered in ascending fashion to represent a random but recovering profile of motor function after acute SCI. We then applied our classification method to the simulated data. The technique successfully classified the data into two classes of what is clearly “delayed” and “advanced” recovery profiles (figure 2B). The results show that, on average, the two classes start at a similar level of motor scores and end at higher but again, similar levels of motor scores. The clear distinction between the two classes is the recovery profiles: one undergoes an initial fast rate of recovery, which slows down at later stages of recovery, and the other group starts the recovery slower but catches up by having relatively faster recovery towards the end of the recovery period.

After checking the validity of our classification to capture profiles of recovery, we applied it to the real motor scores in patients with cervical SCI. Since not all data points were available for all patients, we excluded patients from analysis based on our data cleaning criteria (see methods) (figure 1). An examination of the baseline characteristics of the patients included and excluded from our analysis revealed no meaningful differences (table 1). The patients with less than 10 points total motor score recovery during the 1-year post-injury were classified as “low recovery” group (333 patients). The rest were classified based on the AUNCs to two groups of “delayed recovery” (N=266) and “advanced recovery” (N=307) patients (figures 1&2).

**Table 1.** Baseline characteristics. Data are n (%) unless otherwise stated. SD= Standard Deviation. AIS= ASIA Impairment Scale, where ASIA= American Spinal Injury Association.

|  | Included group (n=906) | Discarded group (n=1306) |
| --- | --- | --- |
| Sex (female) | 165 (18.2%) | 260(19.9%) |
| Mean age at injury in years (SD, range) | 47 (19,13-83) | 50(19,13-88) |
| Severity of initial neurological deficit: |  |  |
| AIS grade A | 131 (14.5 %) | 130 (10.0%) |
| AIS grade B | 61(6.7%) | 50 (3.8%) |
| AIS grade C | 52(5.7%) | 32 (2.4%) |
| AIS grade D | 321 (35.4 %) | 280 (21.4%) |
| No assessment available | 341 (37.6%) | 814 (62.3%) |

**Figure 1.**
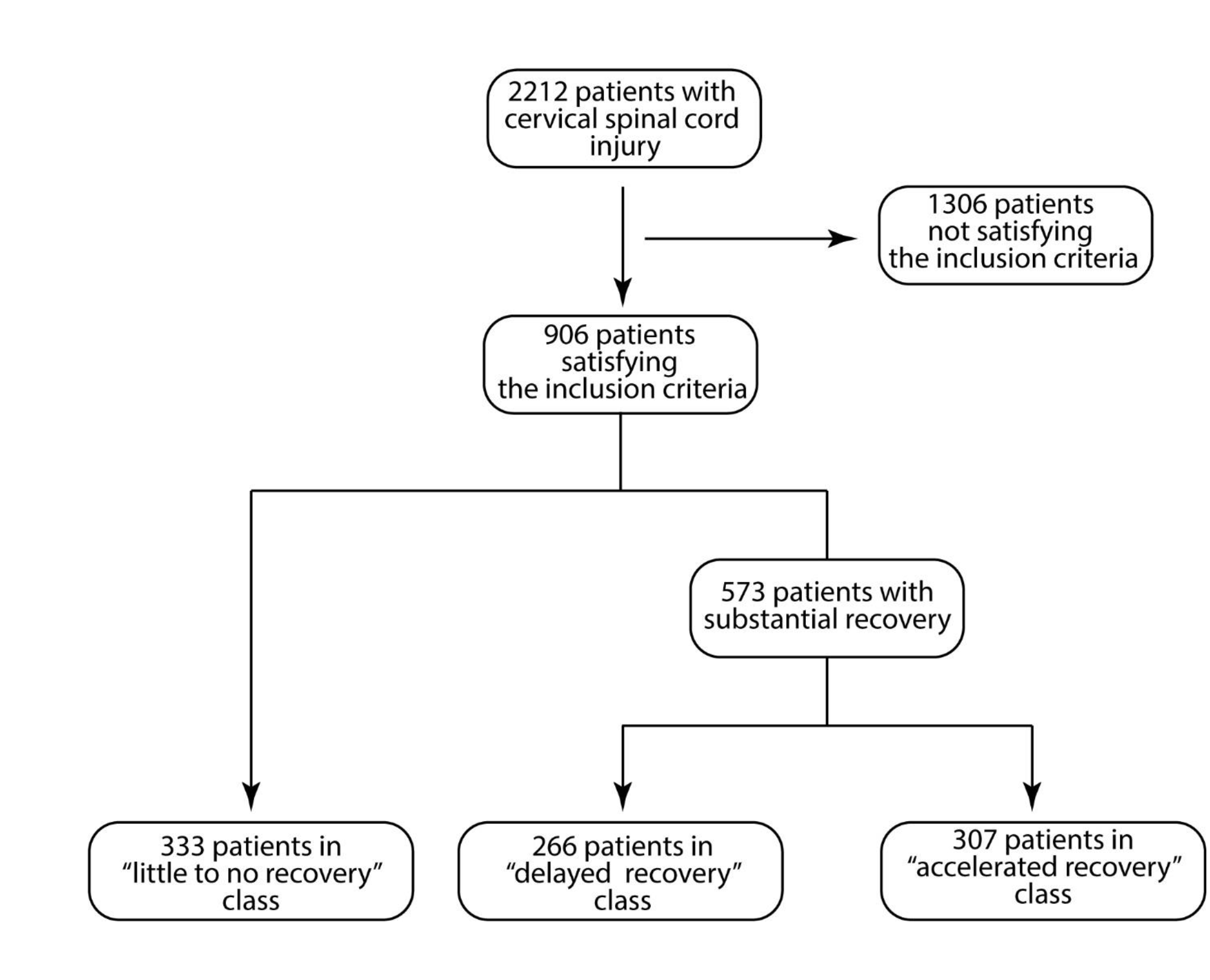
Subject numbers, selection criteria, and subject classification. The inclusion criteria were solely based on the availability of sufficient data points to ensure the possibility of applying the classification algorithm. Patients with two or fewer motor scores throughout their first year of recovery, or missing either of the SCIM or motor score at 12 months were excluded. The remaining patients were classified into three groups based on the profile of neurological recovery using an unsupervised machine learning clustering method.

**Figure 2.**
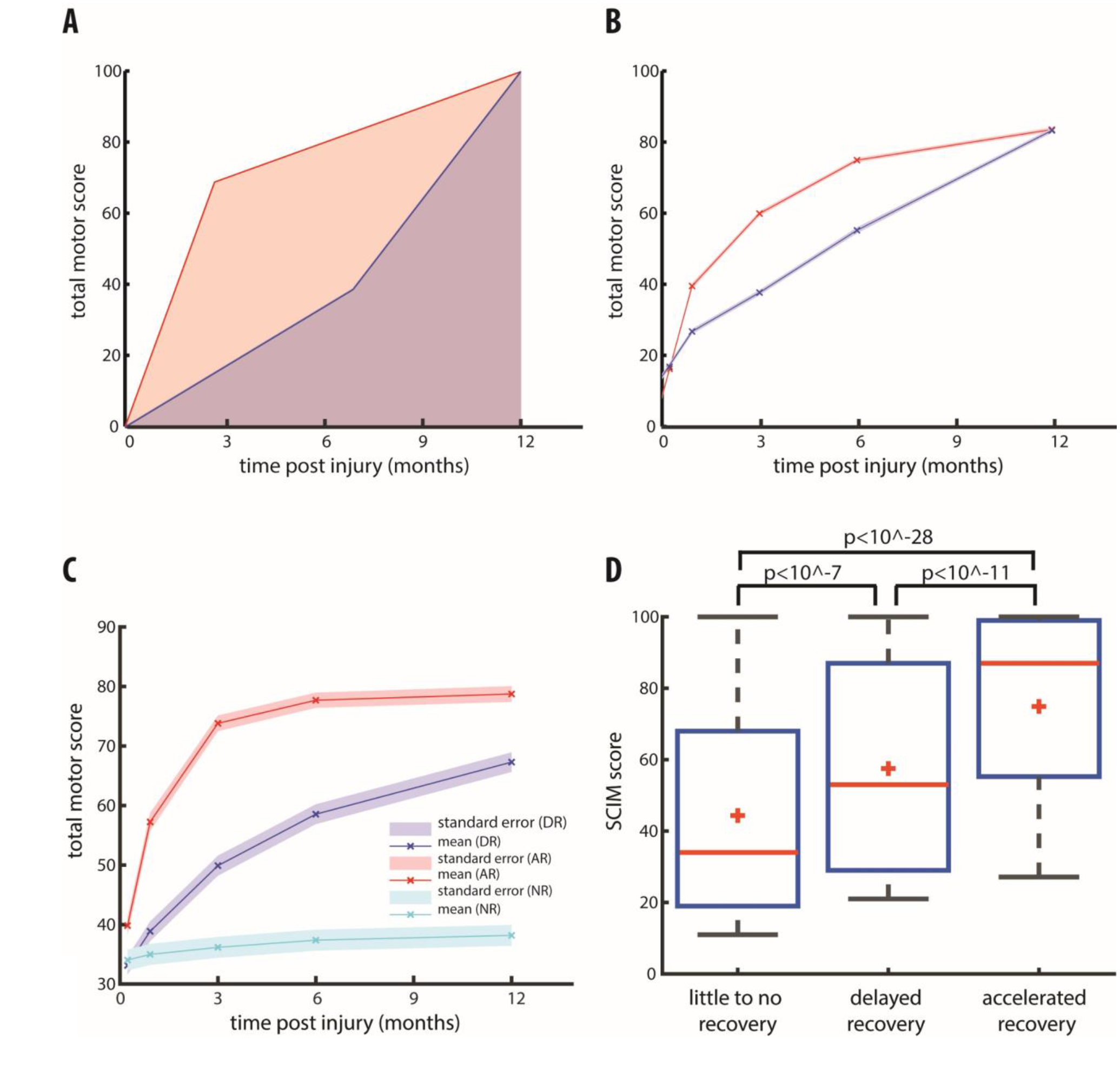
Applying the classification method to simulated and real data. **A**. cartoon depicting the rationale behind the classification method. The advanced recovery profile (red curve) has a larger area under the curve (transparent light red shade) than the delayed recovery profile (transparent light blue shade). Therefore, the area under the recovery curve was used to classify the patients with substantial recovery into delayed and advanced recovery groups. **B**. Applying the classification method to simulated data of substantial neurological recovery successfully classified the simulated patients into advanced recovery (red curve) and delayed recovery (blue curve) groups. **C**. Applying the classification method to real motor scores measured in patients with cervical spinal cord injury. The patients with less than 10 points total motor score recovery during the 1-year post-injury were classified as low recovery group (cyan curve), and the rest were classified into advanced recovery (red curve) and delayed recovery (blue curve) groups. In panels B and C, solid lines represent the estimated mean total motor scores over time, and the light bands represent the standard errors. A cross denotes the mean total motor scores at each measurement time point. **D**. Comparing SCIM scores at 12 months post-injury across low, delayed, and advanced recovery groups. The red horizontal line denotes the median, and the red plus sign displays the mean SCIM scores. Box edges show the 25 and 75 percentiles, and the whiskers display the data range. The p values are computed using the Wilcoxon rank-sum test.

We then compared the SCIM scores at 12 months post injury between the groups (figure 2). The “advanced recovery” group displayed significantly higher SCIM scores (median=87, 25 and 75 percentile=[55,99], range=[27 100]) when compared to both “delayed recovery” (53, [29,87], [21,100], Wilcoxon rank sum test: p<10^-11) and “low recovery “ (34, [19,68], [11,100], p<10^-28) groups. Delayed recovery group also displayed significantly higher SCIM scores than the “low recovery” group (p<10^-7).

These results show that our method can objectively classify the patients to distinct groups based on their profile of neurological recovery with substantial predictive value on the functional outcomes. However, there is a clear difference between these results and the outcome of the simulated data. In the simulated data, the two groups start and end at the same range of motor scores, but in the experimental data, the “advanced recovery” group started and ended at significantly higher levels when compared to the “delayed recovery” group (at week 1: p<10^-11, at week 52: p<10^-8). This means the patients with higher motor scores at the superacute stage have a higher tendency to end up undergoing the “advanced recovery” profile, and the patients in the “advanced recovery” group end up displaying higher motor scores at month 12. While this finding underlines the importance of the neurological recovery profile on the final neurological and functional outcomes, it does not clarify if the recovery profile can directly impact the functional outcome, independent from (or in addition to) the final neurological recovery achieved.

To isolate the effect of neurological recovery profile from the initial and final extent of neurological recovery, we divided our data pool to subgroups of patients that start at the same level (within a range of 10 points) of total motor scores and end at the same level of motor scores after one year. Then, we applied our classification method to each subgroup and finally pooled the “delayed” and “advanced” recovery classes of all subgroups into overall delayed and advanced recovery classes (see methods). Hence, we were able to objectively classify the subjects into two groups where the initial and final motor scores were not significantly different from each other (delayed recovery N_dr_=138, advanced recovery N_ar_=220, Wilcoxon rank-sum test at week 1: p=0.35 & week 52: p=0.10) but displayed two distinct profiles of recovery (figure 3A). Comparing the SCIM scores at month 12, we found that the advanced recovery group displayed significantly higher SCIM scores than the delayed recovery group (advanced recovery: median=80, delayed recovery: median=63, Wilcoxon rank-sum p= 0.0001) (figure 3B). Moreover, we compared the SCIM sub-scores at months 12 among the two groups and all sub-scores were significantly higher in advanced recovery group (subscore1(self-care): median=16(N=209) vs 13(N=149) Wilcoxon rank-sum test p=0.002, subscore2(respiration and sphincter management): median=35(N=202) vs 31(N=156) p=0.006, subscore3(mobility): median=31(N=201) vs 21(N=157) p<10^-4) (figure 3C). Therefore, the rate of neurological recovery, even among patients with a similar initial and final level of neurological recovery, can modulate the functional outcomes. We did not observe any significant difference between the patients in two groups regarding their sex, age, or baseline ASIA Impairment Scale (AIS) grades (figure 3D).

**Figure 3.**
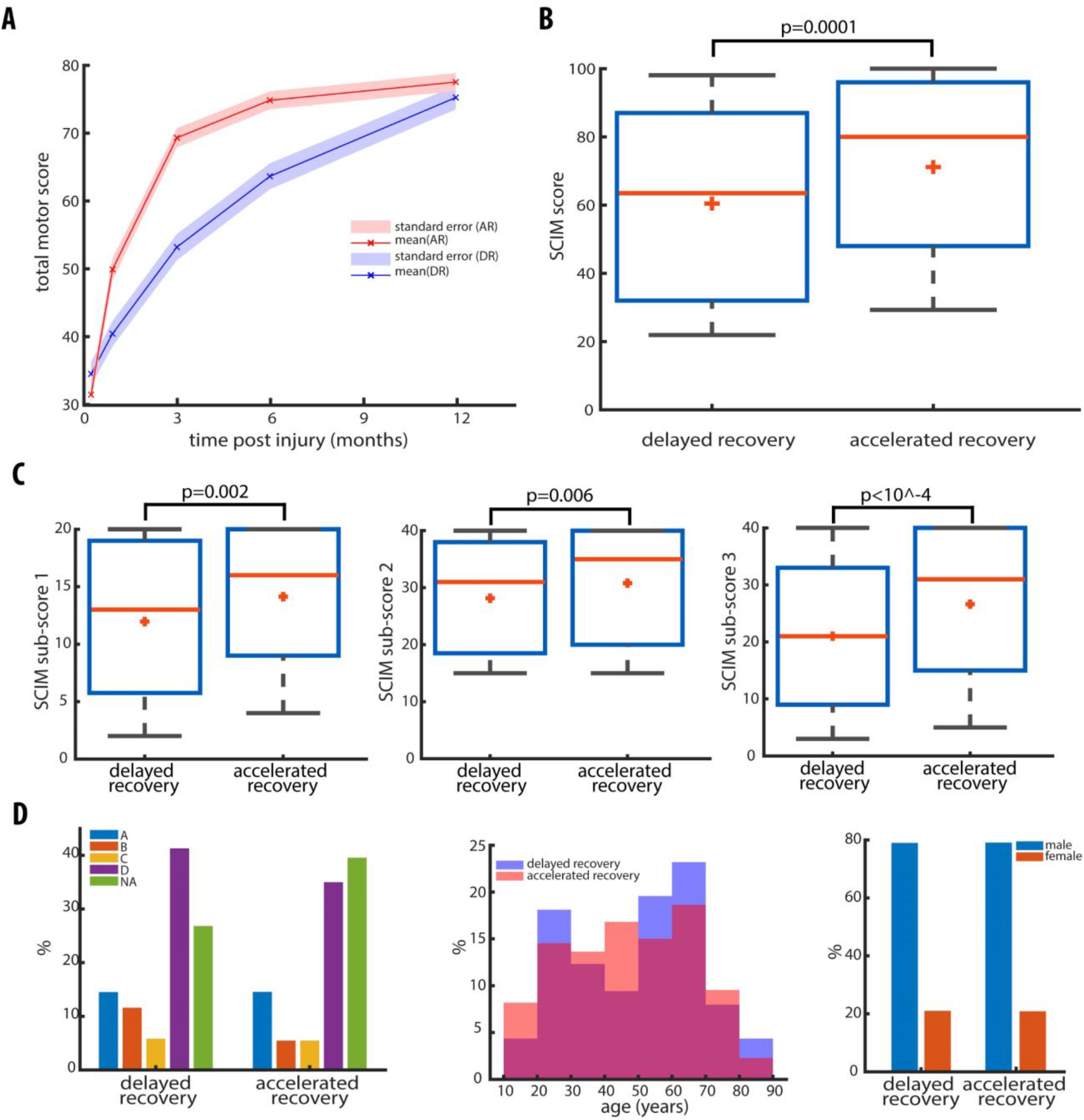
Isolating the temporal profile impact on functional independence. **A**. By imposing targeted restrictions on the initial and final level of total motor scores, we created two groups of delayed (red curve) and advanced recovery (blue curve) that the initial and final motor scores were not significantly different from each other. Solid lines represent the estimated mean total motor scores over time, and the light bands represent the standard errors. A cross denotes the mean total motor scores at each measurement time point. B. Comparing SCIM scores at 12 months post-injury between the two delayed and advanced recovery groups shown in panel A. **C**. All SCIM sub-scores are significantly higher in advanced versus delayed recovery group. Left: self-care, middle: respiration and sphincter management, right: mobility. In panels B and C, the red horizontal line denotes the median, and the red plus sign displays the mean SCIM scores. Box edges show the 25 and 75 percentiles, and the whiskers display the data range. The displayed p values are computed using the Wilcoxon rank-sum test. **D**. Comparing the baseline characteristics between advanced and delayed recovery groups. Left: The distribution of ASIA Impairment Scale (AIS) grades measured at the acute stage in delayed (left set of bars) and advanced (right set of bars) recovery groups. Blue: AIS A, orange: AIS B, yellow: AIS C, purple: AIS D, and green shows the percentage of patients that the AIS grade at the acute stage was not available. Middle: comparing the distribution of patients’ age at admission between delayed (lavender blue) and advanced (pink salmon) recovery groups. The X-axis displays the age at admission. Right: The sex distribution in each recovery group, blue: male and orange: female.

Furthermore, it appears that the neurological recovery is followed by functional independence. Therefore, it can be argued that functional recovery is only slightly delayed in the delayed recovery group. With this logic, one can assume that if we measure SCIM scores at some point later in time, the delayed recovery group will catch up with the advanced recovery group, and we will see the same functional independence across both groups. Since no measurement was done beyond 1-year post-injury, we could not compare the SCIM scores between two groups at a later time point. Instead, we repeated our analysis using only the measurements during the first six months of recovery. This way, we were able to classify the substantial recovery patients to two new groups of “delayed recovery” and “advanced recovery” based on the profiles of recovery during the first six months (figure 4A). Similar to our 12 months analysis, the SCIM scores at six months post-injury for the advanced recovery group was significantly higher than the delayed recovery group [delayed recovery: N_dr_=110, Median =58, advanced recovery: N_ar_= 135, Median=68, Wilcoxon rank-sum test p=0.008](figure 4B). We found that the SCIM scores were still significantly higher at 1-year post-injury in the advanced recovery group [delayed recovery: N1=110, Median =72, advanced recovery: N2= 135, Median=80, Wilcoxon rank-sum test p=0.03] (figure 4C). This result suggests that the effect of the neurological recovery profile on functional independence is robust. While achieving 159 higher functional independence earlier is already a highly desired outcome, the robustness of this effect emphasizes the importance of the neurological recovery profiles.

**Figure 4.**
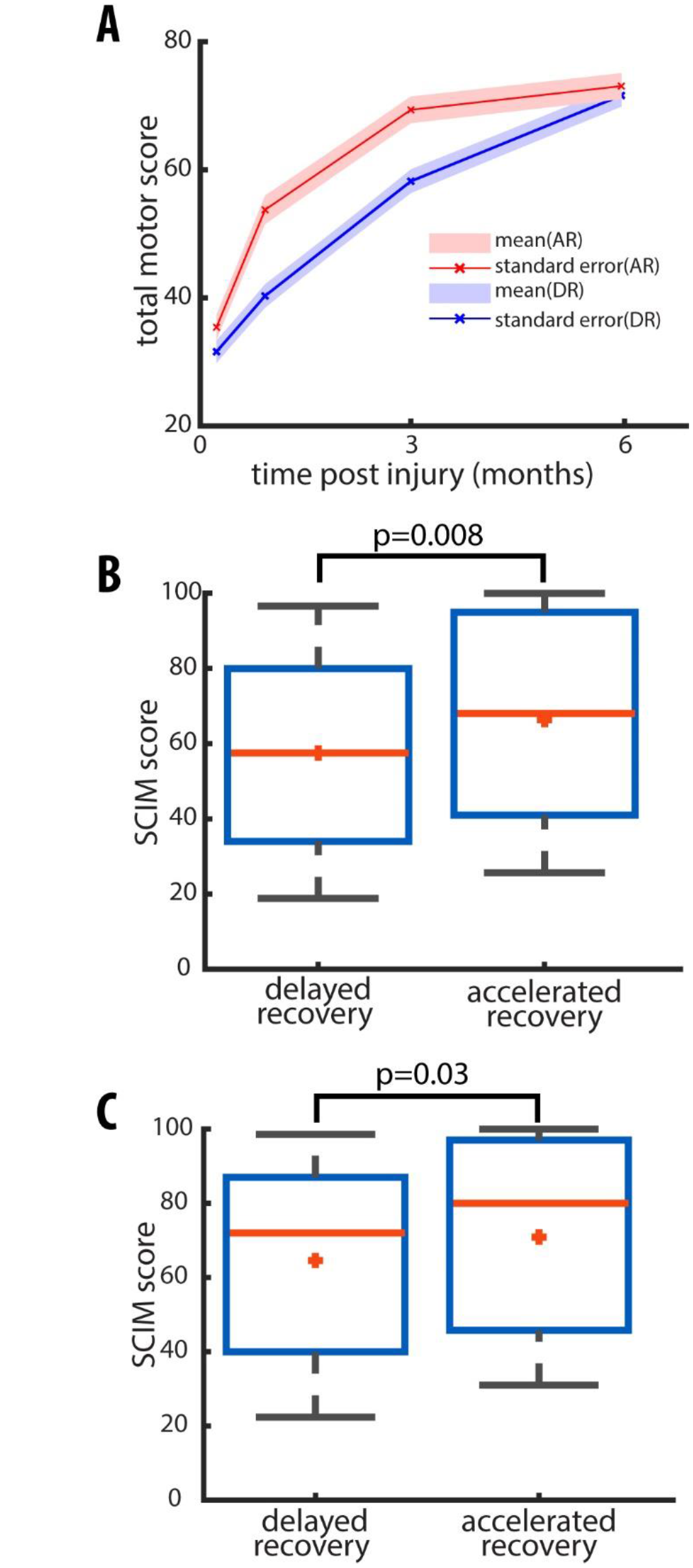
The neurological recovery profile’s impact on functional independence is long-lasting. A. Patients were classified into delayed (blue) and advanced (red) recovery groups based on the six months recovery period. Solid lines represent the mean total motor scores over time, and the light bands the standard errors. **B**. Comparing SCIM scores at six months post-injury between the two delayed and advanced recovery groups. **C**. SCIM scores at 12months post-injury within the same two delayed and advanced recovery groups. In panels B and C, the red horizontal line denotes the median, and the red plus sign displays the mean SCIM scores. Box edges show the 25 and 75 percentiles, and the whiskers display the data range. The displayed p values are computed using the Wilcoxon rank-sum test.

## DISCUSSION

Our analysis aimed to determine the effect of the neurological recovery profile on functional independence after SCI. To do so, we developed an unbiased classification method, which was successful in distinguishing advanced from delayed recovery profile. After adjustment for baseline injury characteristics, advanced neurological recovery to 1 year was associated with higher functional independence compared to slower recovery to the same point. Re-examining changes in total motor scores from acute to 6 months substantiated the functional importance of advanced recovery, and, further, demonstrated that the benefits to independence persisted long-term (i.e., out to 1-year).

There are several possible explanations for why advanced recovery is essential for functional independence. Firstly, there may be a critical window of time after injury, whereby neurological gains can be translated into a meaningful function (e.g., mobility). This can be explained by the injury-induced neuroplasticity^18,19,20,21,22,23,24,25,26^, which is integral to regaining functions^27,28,29,30^, and seems to be limited to the initial weeks to months post injury^31,32,33^. The enhanced neuroplasticity in early stages after injury provides a window of opportunity to learn new strategies in performing tasks geared towards regaining functional independence. Delayed neurological gains can leave patients unable to perform these tasks during the window of opportunity resulting in overall lower functional independence in the long run.

Another potential explanation lies in the timing of rehabilitation. It has been shown that neurotrophic factors are expressed in response to denervation, as well as training^22,23^. Activity-dependent mechanisms can augment plasticity in order to restore useful functional capacities^34,35,36,28,26^. If the rehabilitation efforts are focused during the first few months post-injury, the higher neurological gains can facilitate performing training tasks, which in turn will promote neuroplasticity. This loop re-amplifies the effect of higher neurological gains and results in higher and long-lasting neurological and functional gains.

Regardless of the mechanism underlying the impact of an advanced neurological recovery on functional independence, the presence of such an impact can be used in designing SCI interventional clinical trials. In the planning stages of any clinical trial, there are several critical decisions related to study design that shapes the likelihood of measuring therapeutic effects. In the field of acute SCI, two pressing issues are 1) the selection of outcomes sensitive to detect subtle but meaningful changes in neurological function, and 2) the necessary length of follow-up. Failing to detect the subtle improvements may condemn potentially useful therapies and contribute to inefficiencies in translational research37,38. Also, shortening the duration of a trial (e.g., endpoint examined at six months compared to 1 year) reduces trial costs and the burden of a subject’s participation. Based on our analysis, greater neurological recovery within the first six months in an active treatment group compared to the control group, regardless of whether neurological outcomes continue to recover in the control group and ultimately catch the active treatment group at some later time-point (e.g., no difference at 1-year), is sufficient to infer a functional benefit to the patient. Therefore, the neurological recovery profile as an outcome not only can shorten the required duration of a trial (figure 2 C&D) but also can capture subtle but significant functional improvements missed by final neurological outcome (Figure 3 A&B).

The outcome of the GM1 gangliosides trial in acute spinal cord injury resonates with our observations. While failing to demonstrate efficacy6, the treatment group showed benefits compared to placebo at 4 and 8 weeks post-injury but not at 26 weeks (i.e., trial endpoint). Based on our findings on the impact of advanced neurological recovery, the treatment group may have achieved significantly higher functional independence at 1-year post-injury when compared to the placebo group, if a functional outcome, such as the SCIM, had also been considered. Strikingly similar results were reported for GM1 gangliosides in ischemic stroke39,40, where muscle strength was significantly higher in the treatment group compared to the placebo group at 2 and 4 weeks – differences that were no longer significant at the conclusion of the studies (84 days in one trial and 120 days in another one)39,40. Interestingly enough, patients in the treatment group achieved higher independence in personal care as measured by Barthel Index^41^ when compared to the placebo group^40^, which corresponds with our observations that advanced motor recovery can lead to greater functional returns.

## CONCLUSIONS

Our analysis indicates that the profile of neurological recovery has important long-term effects on functional independence. In particular, advanced recovery promotes functional independence through achieving higher total neurological recovery and facilitating the learning of new strategies during the window of opportunity. It is due to the latter mechanism that the independence benefits remain significant when compared to patients with similar total neurological recovery but delayed recovery profile. For that reason, using the recovery profile as an outcome, not only may shorten the length of clinical trials but can also help better detect subtle improvements, and therefore avoid missing on potentially useful therapies in spinal cord injury. The similarities of findings in SCI and acute ischemic stroke suggests our findings and proposed method may apply to acute ischemic stroke as well.

## METHODS

### Patients

We used data from the European Multicenter study on SCI (EMSCI; http://www.emsci.org). In this project, 21 centers (five centers initially) have gathered a standardized dataset of functional and neurological outcomes of patients with traumatic and ischemic spinal cord injury. The details of various applied treatments were not recorded systematically but ranged from no operative interventions to very early surgical stabilization and decompression of the spinal cord. Following such procedures, patients depending on the level and severity of the injury, participated in various rehabilitation programs. Data for neurological and functional status were collected in 5 time points-within first 15 days, and at months 1, 3, 6, and 12-post-injury. Patients with neuropathy or polyneuropathy, as well as patients who had peripheral nerve lesions not related to the cervical SCI, were excluded from the EMSCI database. The EMSCI study was done in accordance with the ethics standards in the updated version of the 1964 Declaration of Helsinki. The study protocol was approved by the local ethics committees, and the patients gave informed oral consent before entering the study. For this study, we extracted data for all the patients with cervical spinal cord injury.

### Data elements

As a measure of neurological recovery, we used total motor scores from the International Standards for the Neurological Classification of Spinal Cord Injury (ISNCSCI) published by the American Spinal Injury Association (ASIA) ^42^. The total motor score is the sum of all motor scores per 10 paired myotomes (right and left) across the body. Therefore, it provides a single number that tracks overall changes in motor function. We used the Spinal Cord Independence Measure (SCIM) to assess a patient’s functional outcome^43,44^. The SCIM score (0 to 100) was calculated as the sum of the following subscores: self-care (subscore 0 ± 20), respiration and sphincter management (0 ± 40) and mobility (0 ± 40).

### Data Cleaning

Our analysis involved fitting motor scores between time-points to calculate the area under the curve (described in detail below). To ensure accurate model fit, patients were excluded with two or fewer motor scores throughout their first year of recovery. Additionally, any subject with a missing 12-months motor score or SCIM was also excluded. These exclusion criteria were intended to improve the accuracy that the area under the curve reflected how an individual recovered and allow a comparison of neurological and functional outcomes at 12 months.

### Classification method

To explore the neurological recovery profile impact on functional outcomes, we developed a classification method based on the recovery profile. First, we classified subjects with less than ten total motor score points improvement over one year into the “low recovery” group. In the remaining subjects, to estimate the values between the times that motor scores were measured, we used the linear interpolation fit function. In this method, a different linear polynomial is fitted between each data point. Therefore, we were able to have an estimate of the motor score continuously over the full year post-injury. Then, we “normalized” the fitted curves and calculated the area under the “normalized” curves (AUNCs). Based on the size of AUNCs, we classified subjects into “advanced recovery” and “delayed recovery” groups. The logic behind the classification is that if two curves start from the same level and end at a higher motor score at the same level, then the one with an early high recovery rate (“advanced”) will have a higher area under the curve (AUNC) compared to the one with delayed high recovery rate (“delayed”) (figure 2A).

“Normalization” was done to make sure that AUNC is solely influenced by the shape of the recovery curve and not affected by the extent of recovery. To achieve this goal, we subtracted the motor score at week one (estimated by the fitted curve) from the curve and then divided the resulting curve by its value at week 52. In order to objectively classify the curves into small and large AUNCs we used an unsupervised machine learning clustering approach called “K-means”^17^. K-means clustering aims to partition “n” observation into “k” clusters in which each observation belongs to the cluster with the nearest means. Since we are interested in two classes (advanced and delayed recovery), we set the k to 2. We analyzed our data with costume MATLAB (MathWorks, Natick, USA) scripts as well as MATLAB machine learning functions. The K-means clustering algorithm in MATLAB uses randomly generated seeds to determine the starting centroids of the clusters.

### Testing the classification method on simulated data

To test our method of classification, we applied it to a set of simulated post-SCI motor scores (n=1000). For each subject, a vector of 5 random scores between zero and 100 was drawn from a uniform distribution as total motor scores measured at week 1, and at months 1, 3, 6, and 12-post-injury. To mimic the general recovering characteristic of the experimental data, the elements of each vector were ordered in ascending fashion. After classification, the average and standard error of motor score curves for each group were computed and plotted (figure 2B).

### Isolating the effect of the recovery profile

To isolate the direct effect of recovery profile on the functional outcome from the indirect effect, reflected in the motor score level at 1-year post-injury, we systematically divided the recovering patients into ten groups. Each group consisted of patients with starting (super acute) motor scores bounded to specific ranges (MS_a_ ∈ [0,10],(10,30],(30,50], or [50,70]) and final (1-year post-injury) motor score bounded to a higher defined range (MS_1y_ ∈ [20,40],(40,60],(60,80], or [80,100]). Applying our classification method to each group, we classified them to 10 “advanced” and 10 “delayed” recovery subgroups. By pooling “advanced” and “delayed” recovery subgroups, we generated two overall classes of “advanced” and “delayed” recovery that, on average, display similar motor score levels at the start and end of the recovery period (figure 3A).

### Six-months recovery classification

To test the robustness of the recovery profiles effect on functional outcomes, we applied our classification method to the motor scores measured during the first six months post-injury instead of 12 months. Then, we compared the SCIM scores at months 6^th^ and 12^th^ between the two new “delayed” and “advanced” recovery groups and observed the progression of difference from month 6^th^ to month 12^th^. For this analysis, patients with two or fewer motor scores available during the six months post-injury, as well as patients missing either of SCIM score measurements at months 6^th^ and 12^th^, were excluded.

## Data Availability

The data that support the findings of this study are available from the corresponding author upon reasonable request. Quantification and statistical analysis were performed in Matlab, and the code will be provided upon request.

https://www.emsci.org/index.php/project/the-database

## AUTHOR CONTRIBUTIONS

Conceptualization, N.K. and J.K.; Data collection, R.A., L.G., Y-B.K., D.M., R.R., N.W., and A.C.; Formal analysis, N.K.; Investigation, N.K. and J.K.; Methodology, N.K. and J.K.; Software, N.K.; Writing - Original Draft, N.K.; Writing - Review and Editing, N.K., R.A., L.G., Y-B.K., D.M., R.R., N.W., A.C., and J.K.

## DATA AND SOFTWARE AVAILABILITY

Quantification and statistical analysis were performed in Matlab, and the code will be provided upon request.

